# Delay and Probability Discounting of HIV Vaccines*

**DOI:** 10.1101/2025.03.27.25324754

**Authors:** Promise Tewogbola, Victoria Bodunde, Oluwakamikun Adekunle, Deborah Odaudu

## Abstract

This study investigated how delay and probability discounting impact the perceived value of hypothetical HIV vaccines under different monetary cost conditions. Using hypothetical purchase tasks, two experiments assessed how sexual and gender minorities (SGMs) made trade-offs between immediate versus delayed vaccine access and certain versus uncertain availability. In Experiment 1 (*N* = 260), participants chose between a 60% effective vaccine available immediately and a 99% effective vaccine with varying delays. Experiment 2 (*N* = 246) involved a choice between a 55% effective vaccine with certain availability and a 95% effective vaccine with uncertain availability. In both experiments, participants were randomly assigned to conditions where the vaccine was either free or available for a $150 fee. Hyperbolic discounting models effectively described delay (*R*^2^ = 0.96 − 0.99) and probability discounting (*R*^2^ = 0.95 − 0.99). While vaccine cost did not significantly affect discounting, individual characteristics emerged as significant predictors. Age (*β* = 0.01, *p <* 0.05) and prior STI testing (*β* = −0.19, *p <* 0.05) influenced delay discounting, while probability discounting was influenced by age (*β* = 0.01, *p <* 0.05), identifying as gay or bisexual (*β* = −0.15, *p <* 0.05), health insurance status (*β* = 0.33, *p <* 0.05), prior STI testing (*β* = 0.18, *p <* 0.05), and political liberalism (*β* = −0.004, *p* = 0.05). This research is among the first to model HIV vaccine acceptance using behavioral economic methodology. When designing vaccine distribution programs, public health strategies may benefit from considering factors such as age, STI testing behaviors, health insurance, and political views. Future research should explore more vulnerable populations such as sex workers and people who inject drugs (PWIDs), and investigate how the perceived value of HIV vaccines may intersect with other health interventions.

## 1 Introduction

In the United States, HIV remains a public health issue, despite the country spending approximately 17% of its GDP on health-related expenses, according to the World Bank’s Global Health Expenditure database (World Bank, 2024). In impoverished urban areas, HIV prevalence mirrors that of several low-income countries experiencing widespread epidemics (Cohen, 2018; Denning & DiNenno, 2010). Current prevention and treatment strategies in the U.S. face substantial obstacles. For instance, despite the recommendations for regular HIV testing from public health organizations such as the Centers for Disease Control and Prevention (CDC) and the United States Preventive Services Task Force (USPSTF) for at-risk populations, approximately 15% of those infected are still unaware of their HIV-positive status. This gap in awareness has significant public health implications, as this group accounts for 40% of new HIV transmissions (DiNenno et al., 2017; Li et al., 2019; Owens et al., 2019).

While prophylactic drugs such as pre-exposure prophylaxis (PrEP) have been shown to reduce HIV transmission, only 36% of eligible individuals in the U.S. receive them (CDC, 2023). This limited coverage is further complicated by significant racial disparities in access and treatment outcomes, particularly among Black and Hispanic populations who are disproportionately affected by HIV (CDC, 2021). Moreover, maintaining long-term daily adherence to these prophylactic drugs presents a major challenge (CDC, 2022; HIVInfo, 2021; Lyons et al., 2021; Oh et al., 2009). Poor adherence can lead to drug-resistant HIV strains, potentially rendering current antiretroviral therapies (ARTs) and pre-exposure prophylaxis (PrEP) ineffective, thus increasing HIV/AIDS mortality rates (Apetroaei et al., 2024). These multiple challenges highlight the limitations of current prevention approaches.

Given these challenges, developing a safe and effective HIV vaccine could be transformative in reducing HIV infections both in the US and globally. Vaccines have historically been the most efficient method for preventing and eradicating infectious diseases (Ellenberg & Chen, 1997; Habersaat & Jackson, 2020; Prüß, 2021). Success stories with smallpox, diphtheria, pertussis, tetanus, typhoid, rabies, measles, polio, and more recently, COVID-19 demonstrate how preventive vaccines can provide long-lasting protection and community immunity while eliminating many challenges associated with treatment-based approaches, including high costs, delayed onset of treatment, drug resistance, adverse side effects, poor adherence, and stigma (Duncan et al., 2012; Dybul et al., 2021; Havlir et al., 2020; Johnson & Neilands, 2007; Van Tam et al., 2011). Recent developments in mRNA technology, spurred by the success of COVID-19 vaccines, have reignited hope for an HIV vaccine. In March 2022, for instance, the National Institute of Allergy and Infectious Diseases (NIAID) initiated clinical trials for three experimental HIV vaccines using mRNA technology derived from approved COVID-19 vaccines (Rogers, 2022). As Dr. Anthony Fauci, then director of NIAID, noted, *“HIV research absolutely helped COVID-19. Now that we have a successful vaccine with mRNA, it’s going to go back. Everything that goes around comes around. We’re going to hopefully get more insight into HIV vaccines*.*”* (Stulpin, 2021).

That said, the eventual success of an HIV vaccine program will depend not only on vaccine development but also on understanding and addressing factors that influence vaccine acceptance among high-risk populations. To understand and predict how individuals might approach decisions about future HIV vaccines, behavioral economics offers valuable theoretical frameworks, particularly through the concepts of delay and probability discounting. These approaches can help model how members of high-risk groups evaluate and make decisions about accepting an HIV vaccine, especially when faced with various uncertainties and delays in vaccine availability.

Discounting refers to how the perceived value of a commodity decreases as a function of various factors, such as temporal distance or uncertainty of access (Mazur, 1987; Rachlin, 2000). This concept is particularly relevant to HIV vaccine acceptance, as individuals must weigh immediate versus delayed benefits, as well as certain versus uncertain outcomes. In healthcare decision-making, people often prefer immediate, concrete outcomes over delayed, abstract benefits – even when the delayed option might provide greater long-term protection. This tendency can significantly impact preventive health behaviors, including vaccine acceptance.

Two forms of discounting are especially pertinent to HIV vaccine acceptance: delay discounting and probability discounting. Delay discounting refers to the devaluation of a commodity as the time to its receipt increases. In contrast, probability discounting describes the devaluation of a commodity based on the uncertainty of its occurrence (Rachlin, 2006). To quantify how individuals evaluate delayed or uncertain access to HIV vaccines, several mathematical models of discounting can be applied. These models provide frameworks for understanding and predicting vaccine acceptance decisions under various conditions.The most widely applied model is the hyperbolic discounting function, given as:

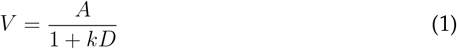

where *V* is the subjective value of the delayed reward, *A* is the amount of the delayed reward, *D* is the delay, and *k* represents the individual’s discounting rate. In some analyses, the parameter *k* serves as a key indicator – lower *k* values suggest a greater willingness to wait for the vaccine (self-control), while higher *k* values indicate a stronger preference for immediate alternatives (impulsivity). This model effectively captures how people often demonstrate time-inconsistent preferences, particularly relevant in healthcare decisions where benefits may be delayed (Ainslie, 1975). Extended versions of this model introduce an additional parameter (*s*) to enhance sensitivity to delay:

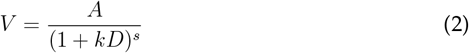

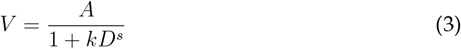

where a higher value of *s* indicates a greater sensitivity to delay.

Probability discounting follows a similar hyperbolic model, with subjective value *V* declining as the odds against accessing that commodity (*θ*) increase:

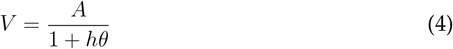

where *h* represents the discounting rate with lower values indicating risk tolerance and higher values reflecting risk aversion (Rachlin et al., 1991). Rachlin (2006) also describes the two-parameter hyperboloid forms of probability discounting:

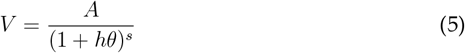

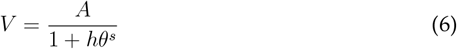

A complementary method for quantifying discounting is the Area Under the Curve (AUC) approach, which provides a nonparametric measure of discounting severity (Myerson et al., 2001):

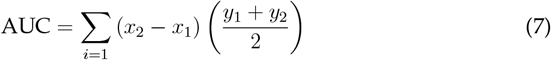

where *x*_1_ and *x*_2_ are normalized delay/probability values, and *y*_1_ and *y*_1_ are the corresponding normalized indifference points. Higher AUC values indicate less discounting (greater willingness to wait or accept uncertainty), whereas lower AUC values indicate more impulsive decision-making.

Despite advances in HIV prevention strategies and promising developments in vaccine technology, significant gaps remain in understanding how potential recipients will evaluate and make decisions about HIV vaccine acceptance. Behavioral economic principles have shown promise in influencing a range of health behaviors, such as alcohol use (Vuchinich & Simpson, 1998), smoking (Mitchell, 1999), drug use (Monterosso et al., 2007), and vaccination decisions (Halilova et al., 2022). However, their application to HIV vaccine acceptance has not been extensively studied. We particularly lack knowledge on how different access conditions - such as monetary costs, timing, and certainty of availability - might impact vaccine acceptance among high-risk populations.

Consequently, the present study has three aims: (1) to evaluate the utility of delay and probability discounting curves, generated through Simulated Purchase Tasks (SPTs), in describing how the perceived value of HIV vaccines decays with increasing delays and uncertainty of access. This approach will allow the quantification of how individuals trade off immediate versus delayed vaccine access, as well as certain versus uncertain availability; (2) to evaluate how contextual factors influence HIV vaccine acceptance. Specifically, the influence of vaccine cost (free versus paid), timing of availability (immediate versus delayed), and certainty of access (varying levels of probability) on individuals’ willingness to accept vaccination will be evaluated; (3) to evaluate how discounting of HIV vaccines vary based on the personal and interpersonal characteristics of individuals (age, gender, ethnicity, socioeconomic status, history of HIV risk behavior, religious and political orientations).

## 2 Experiment 1

The purpose of Experiment 1 was to examine the impact of the cost of delayed HIV vaccines — available either for free or for a $150 fee - on how their value was discounted. Participants were randomly divided into two groups: one with access to free HIV vaccines and the other with the option to purchase the vaccines for $150. In both conditions, participants made choices between a less effective vaccine that was available immediately and a more effective vaccine available after a series of increasingly longer delays.

### 2.1 Participants

A sample of 393 participants was recruited through Reddit and organizations serving Sexual and Gender Minorities (SGMs) throughout the United States. The study protocol was approved by the Institutional Review Board (IRB) of a Midwestern state university. The survey, hosted on Qualtrics, presented participants with an informed consent document, and only those who provided consent could proceed with the study. Participation was voluntary and uncompensated.

### 2.2 Procedure

The following measures were used in Experiment 1. Note that while demographic information and personal covariates are listed here for contextual relevance, they were collected after the completion of both Experiment 1 and Experiment 2.

> Participants were permitted to partake in a simulated HIV Vaccine Delay Discounting Task (HVDDT) where they were provided with the following instructions:
>
> Please read and consider the following. Imagine a situation in which two different HIV vaccines were available for XX for anybody who wanted to get immunized. HIV Vaccine A is 99% effective but only available in stock after a certain delay. HIV Vaccine B is 60% effective and is immediately available. Both vaccines are easy to use and take less than 5 minutes to administer. The following questions ask about whether or not you would get an HIV vaccine for XX, given a series of delays in receiving the vaccine itself. Remember that there are no “right” or “wrong” answers. Please respond honestly, as if you were actually in this scenario.

The value of XX characterized the experimental condition the participants were assigned to – whether HIV vaccines are available for free or for a $150 fee. Participants reported their preferences between an immediately available but less effective HIV Vaccine B and a more effective HIV Vaccine A only available after a series of delays. The specific delays to assessing the more effective HIV Vaccine B were: 1 day, 1 week, 1 month, 6 months, 1 year, and 5 years.

#### Risk Behavior Assessment

Participants responded to a series of dichotomous yes-no questions designed to assess their risk of contracting HIV. These include previous HIV testing, recent STI testing (excluding HIV), PrEP usage in the past year, history of injecting non-prescribed drugs, condomless sex in the past year, and non-prescribed drug use in the past 30 days.

#### Subjective HIV Risk Perception

Participants rated their perceived risk of HIV infection on a VAS ranging from 0 (“Very unlikely”) to 100 (“Very likely”) in response to the question: *“What is your gut feeling about how likely you are to get infected with HIV?”*

#### Trust

Trust in scientific research, medical professionals, and the US government were each assessed on a 100-point scale (0 = no trust at all; 100 = a great deal of trust).

#### Political Views

Participants rated their political views on a VAS ranging from 0 (*“Very conservative/not liberal at all”*) to 100 (*“Very liberal/not conservative at all”*).

#### Discrimination Experience

Participants indicated (*Yes/No*) whether they had experienced discrimination based on race, gender, or sexual orientation.

#### Vaccination History

Participants reported (*Yes/No*) whether they had received any vaccine (e.g., flu, COVID-19, etc.) in the past 3 years.

#### Health Insurance Coverage

Participants responded *“Yes”* or *“No”* to the question:

*“Do you currently have health insurance or health care coverage?”*

#### Demographics

Participants provided comprehensive demographic information as part of the study. They reported their assigned gender at birth, choosing from options including *‘Male’, ‘Female’, ‘Other’*, or *‘Prefer not to say’*. Regarding their sexual partners in the past year, participants selected from *‘Men only’, ‘Women only’, ‘Both men and women’, ‘I have not had sex’*, or *‘Other’*. Age was reported in years. For race and ethnicity, participants self-identified t heir r acial identity, with options including *‘Black/African American’, ‘Hispanic/Latino’, ‘Indigenous Peoples’, ‘Middle Eastern or North African’, ‘Pacific Islander’, ‘White or Caucasian’, ‘Asian’*, or *‘Other’*. Marital status was categorized as *‘Married’, ‘Living together as married’, ‘Separated’, ‘Divorced’, ‘Widowed’*, or *‘Never married’*. Total annual household income was reported in brackets: *‘$0-$9*,*999’, ‘$10*,*000-$24*,*999’, ‘$30*,*000-$49*,*999’, ‘$50*,*000-$74*,*999’, ‘$75*,*000-$99*,*999’, ‘$100*,*000-$149*,*999’*, or *‘$150*,*000+’*. Educational attainment was assessed by highest level completed, ranging from *‘Never attended school’* to *‘Any post-graduate studies’*, with intermediate options including *‘Grade 12 or GED’, ‘Some college, Associate’s Degree, or Technical Degree’*, and *‘Bachelor’s Degree’*. Participants also described their residential area as *‘An inner-city area’, ‘A suburban area’, ‘A town’, ‘A village’*, or *‘Rural or countryside’*.

### 2.3 Statistical Analyses

The two-stage approach described by Kaplan and colleagues (2021) was used to evaluate the discounting of both delayed HIV vaccines and monetary rewards. First, individual-specific discounting rates were estimated using the AUC method. The process involves normalizing both delays (D) and the value of the more effective HIV vaccine (A), such that they span from 0 to 1. AUC is then determined by summing the areas of the trapezoids formed between each interval, calculated using the following:

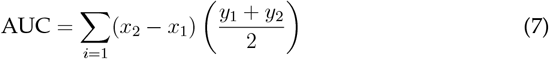

where *x*_1_ represents the normalized smaller delay to accessing the more effective HIV vaccine, *x*_2_ is the normalized larger delay, and *y*_1_ and *y*_2_ are the corresponding normalized valuations of the more effective HIV vaccine. That said, the one-parameter hyperbolic discounting model was also used to describe delay discounting at the aggregate level:

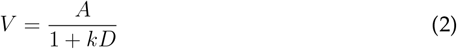

where *V* is the subjective value of the delayed vaccine; *A* is the amount or magnitude of the delayed vaccine; *D* is the delay in assessing the more effective vaccine; and, a parameter, *k*, reflects the discounting rate.

The primary outcome measures of interest in this study were the AUC values for discounting delayed HIV vaccines, which were obtained from the first stage of data analysis. Due to the non-normal distribution of AUC values, a logarithmic transformation was applied before proceeding to the second stage of analysis. This subsequent phase involved a multiple linear regression analysis conducted through three separate models. The first model used the experimental conditions as predictors to assess their direct impact on discounting AUC values. The second model examined the influence of demographic variables and other personal characteristics of participants. The third model combined both sets of predictors, offering a more holistic view of the factors affecting the discounting indices.

Several categorical variables were included in the analysis, each assigned specific coding schemes. Sex at birth was classified as ‘Male’ (coded as ‘1’) versus all other categories (coded as ‘0’). The biological sex of sexual partners was categorized as exclusively ‘Men’ or ‘Both men and women’ (coded as ‘1’), with all other responses grouped together. Ethnicity was dichotomized as ‘White’ (coded as ‘1’) versus other ethnicities. Socioeconomic status was based on household income, with those earning below $75,000 coded as ‘1’ and those earning above this threshold coded as ‘0’. Marital status was distinguished between individuals who were ‘Married’ or ‘Living together as married’ (coded as ‘1’) and those with other relationship statuses. Educational attainment was grouped into ‘Some college or higher’ (coded as ‘1’) versus lower levels of education. Residential location was categorized as ‘Inner city’ or ‘Suburban’ (coded as ‘1’), with all other areas classified separately.

For behavioral and health-related variables, negative responses (‘No’) served as reference categories throughout the analysis. This also applied to HIV testing history, other STI screening, PrEP usage, injection drug use, experiences of racial discrimination, health insurance status, unprotected sexual activity, substance use, and vaccination history. Continuous variables retained their original scaling, including age, trust (in science, in government, and in medical professionals), religiosity, political liberalism, and perceived HIV risk, where higher scores indicated greater levels of each measure.

### 2.4 Results

Incomplete or missing entries were identified and excluded from further analysis. As a result, 133 participants (33.84% of the dataset) were removed based on this exclusion criterion. Consequently, data from 260 participants met the inclusion criteria and were retained for the second stage of analysis. A summary of the demographic characteristics of the final sample is presented in Table 1.

**Table 1.** Characteristics of Participants Meeting Inclusion Criteria for Experiment 1 (N = 260)

|  | Mean (SD); N (%) |
| --- | --- |
| Vaccine Cost |  |
| Free | 127 (48.85%) |
| \$150 | 133 (51.15%) |
| Demographics and personal characteristics |  |
| Age | 29.37 (7.86) |
| Born male | 80 (28.07%) |
| Bisexual/Had sex with other men | 100 (35.09%) |
| White | 144 (50.18%) |
| Married/Living together as married | 55 (19.3%) |
| Household income <\$75000 | 118 (41.4%) |
| At least some college education | 180 (63.16%) |
| Done HIV test | 134 (47.02%) |
| Done other STI test | 89 (31.23%) |
| Taken PrEP | 39 (13.68%) |
| Injected drugs | 15 (5.26%) |
| Been racially discriminated | 142 (49.82%) |
| Has health insurance | 183 (64.21%) |
| Had unprotected sex | 117 (41.05%) |
| Consumed alcohol | 140 (49.12%) |
| Consumed other drugs | 76 (26.67%) |
| Taken other vaccine | 175 (61.4%) |
| Inner city/Suburban residency | 145 (50.88%) |
| Trust in science | 86.15 (19.19) |
| Trust in medical professionals | 76.87 (23.27) |
| Trust in government | 46.34 (28.87) |
| Liberalism | 84.49 (22.43) |
| Religiosity | 28.39 (29.78) |
| Perceived risk of HIV infection | 19.80 (24.19) |

Note. For discrete variables, the number of participants and percentage of sample are included. For continuous variables, mean and SD are included.

The hyperbolic discounting function effectively modeled delay discounting of HIV vaccines in the contexts of both the free and the $150 fee conditions, demonstrating high accuracy. This is evidenced by the substantial R-squared values obtained for the two experimental groups (Free: *R*^*2*^ = 0.99; $150 fee: *R*^*2*^ = 0.96). The computed aggregate k-value was 0.0010 for the free condition and 0.0014 for the $150 fee condition. Figure 1 displays the optimally fitted hyperbolic discounting function for each of these conditions. Furthermore, no significant differences were found in the AUC between the two experimental groups (Free: *M* = 0.60, *SD* = 0.35; $150 fee: *M* = 0.53, *SD* = 0.38; *t*(258) = 1.52, *p* = 0.13). The comparative AUCs for both conditions are depicted in a bar chart in Figure 2. Moreover, when examining the correlation between the log-transformed AUCs from the Monetary Delay Discounting Task and the HIV Vaccine Delay Discounting Task, it was found to be not significant, *r*(205) = 0.02, *p* = 0.75. Figure 1 displays the aggregate discounting curves for delayed HIV vaccines and delayed monetary rewards.

**Figure 1.**
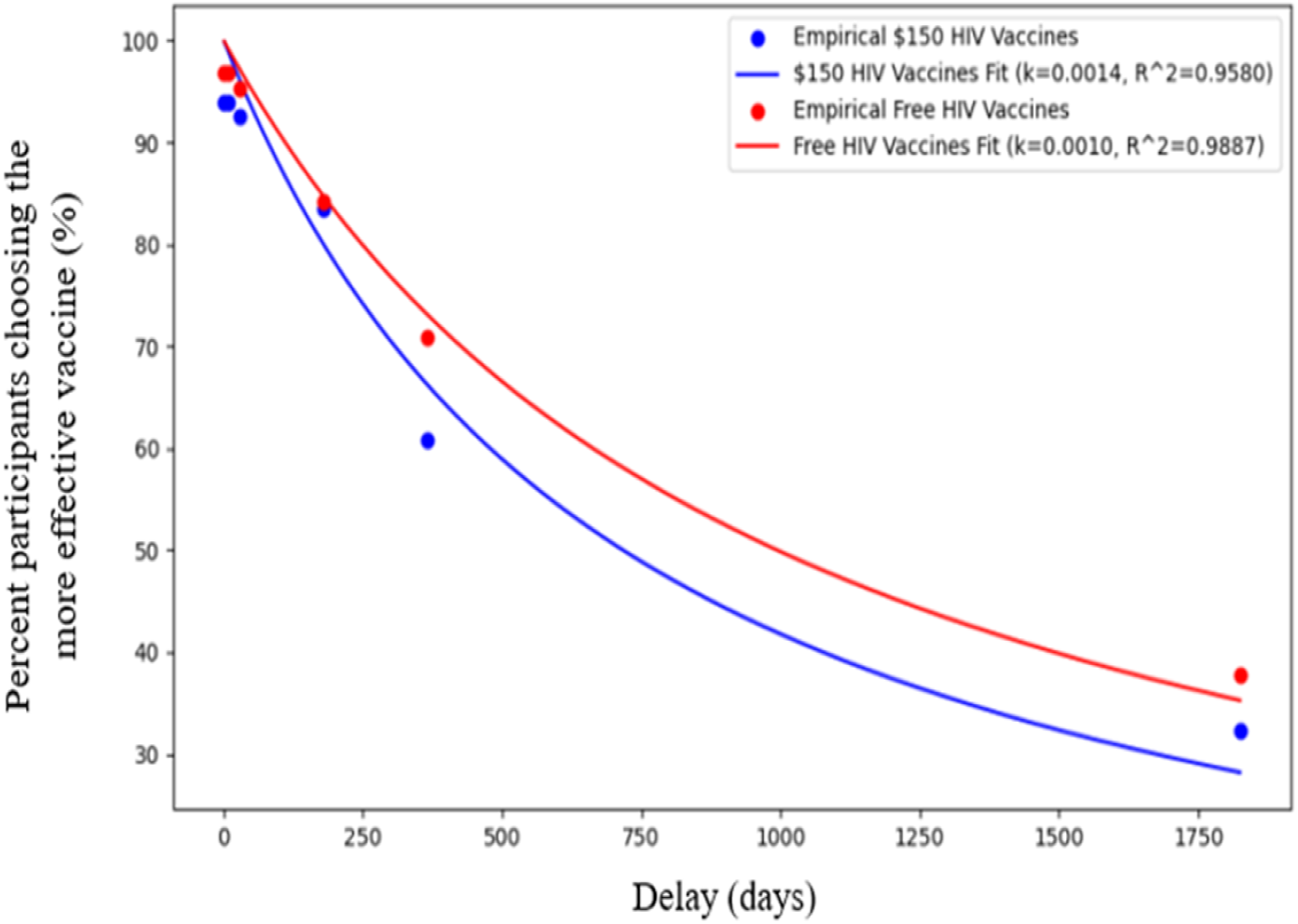
Aggregate Delay Discounting of a More Effective but Delayed HIV Vaccines Available for Free vs for $150. *Note*. Plotted are group discounting curves for a more effective but delayed HIV vaccine available for free (red) and for a $150 fee (blue). Curves were plotted using the one-parameter hyperbolic discounting function.

**Figure 2.**
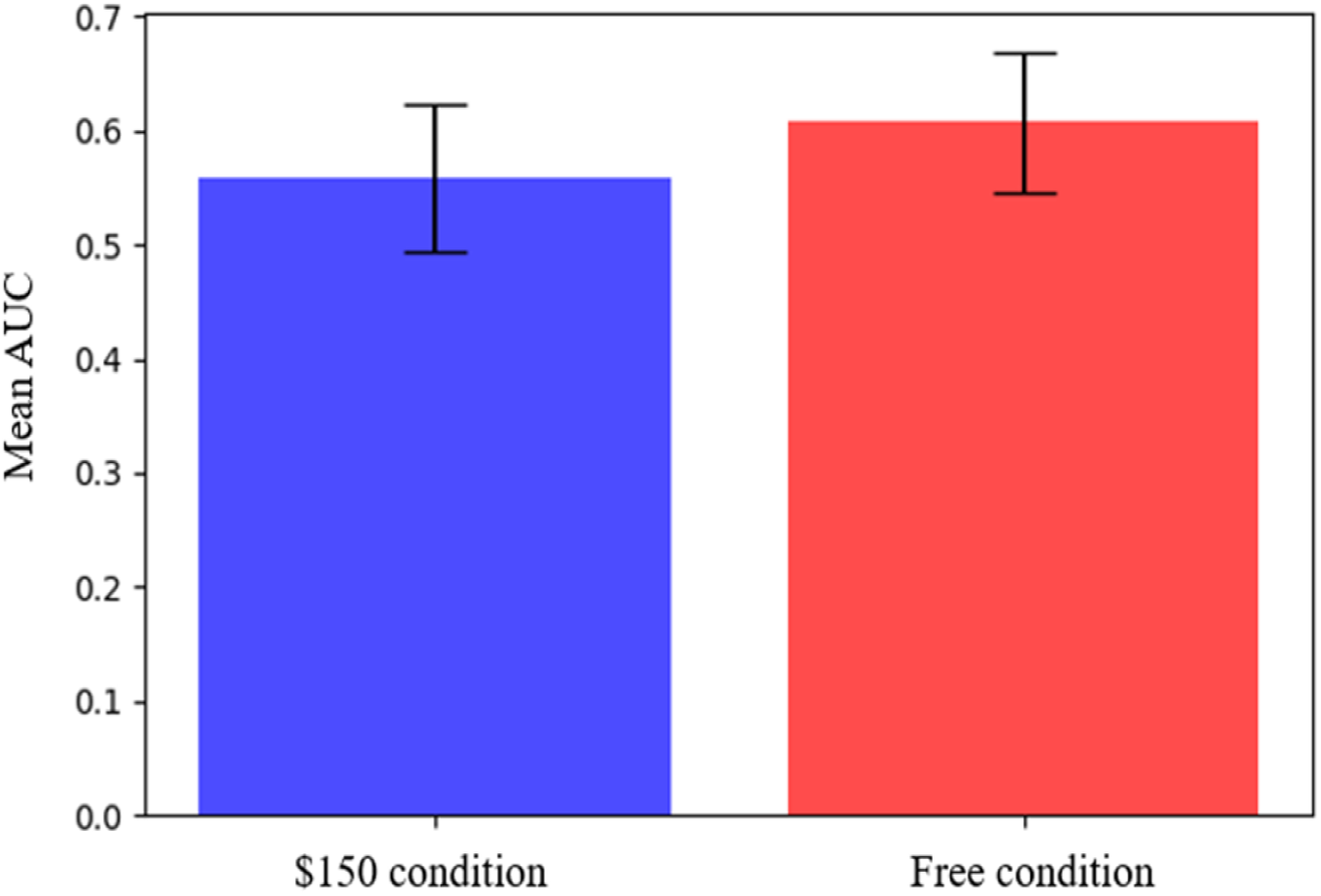
Bar Charts of Mean AUCs for a More Effective but Delayed HIV Vaccines Available for Free vs for $150. Note. The bars represent the group mean AUCs for a more effective, delayed HIV vaccines available for free (red) and for a $150 fee (blue). Error bars indicate 95% confidence intervals.

Results from three regression models investigating the determinants of variability in the log-transformed delay discounting AUCs are presented in Table 2. The first model, which considered the experimental context of HIV vaccines being available either for free or at a $150 fee as predictors, found that the effect size of this context on delay discounting for HIV vaccines was small (*R*^*2*^ = 0.004). There were no significant differences between the reference comparison group (vaccines for free) and the other condition (vaccines for $150), despite the mean AUC being greater for the reference group. In contrast, the second model, which focused on demographics and other personal characteristics of participants, offered more explanatory power (*R*^*2*^ = 0.159). Within this model, the significant predictors of delay discounting included a history of undergoing screening tests for STIs other than HIV (*β* = −0.19, *p* < 0.05) and age (*β* = 0.01, *p* < 0.05).The third model, integrating both the experimental context and personal factors, demonstrated a marginal increase in explanatory power over the second model (*R*^*2*^ = 0.16).

**Table 2.** Regression Results Showing Effect of the Experimental Context of HIV Vaccines Available either for Free or at a $150 Fee and Personal Characteristics on Delay Discounting AUCs.

| Predictor | log AUC |  |  |
| --- | --- | --- | --- |
| | $\beta$ | $p$ | $r^2$ |
| Model 1: Experimental context |  |  |  |
| Vaccines for free (reference) | **-0.37** | **< 0.001\*\*\*** | 0.004 |
| Vaccines for \$150 fee | -0.06 | 0.31 | |
| Model 2: Personal characteristics |  |  |  |
| Age | **0.01** | **0.02\*** | 0.159 |
| Born male | -0.02 | 0.85 |  |
| Bisexual/Had sex with other men | 0.09 | 0.28 |  |
| White | -0.11 | 0.21 |  |
| Married/Living together as married | -0.08 | 0.40 |  |
| Household income <\$75000 | 0.09 | 0.26 | |
| Education |  |  |  |
| Done HIV test | 0.13 | 0.13 |  |
| Done other STI test | **-0.19** | **0.03** |  |
| Taken PrEP | -0.09 | 0.46 |  |
| Injected drugs | -0.31 | 0.08 |  |
| Been racially discriminated | 0.14 | 0.12 |  |
| Has health insurance | -0.17 | 0.28 |  |
| Had unprotected sex | 0.01 | 0.91 |  |
| Consumed alcohol | -0.01 | 0.90 |  |
| Consumed other drugs | 0.05 | 0.51 |  |
| Taken other vaccine | -0.04 | 0.80 |  |
| Inner city/Suburban residency | 0.09 | 0.30 |  |
| Trust in science | 0.0002 | 0.94 |  |
| Trust in medical professionals | 0.0001 | 0.54 |  |
| Trust in government | 0.0005 | 0.78 |  |
| Liberalism | -0.003 | 0.21 |  |
| Religiosity | 0.002 | 0.24 |  |
| Perceived risk of HIV infection | 0.002 | 0.30 |  |
| Model 3: Experimental context and personal characteristics |  |  |  |
| Predictors in Model 1 and 2 | YES | YES | 0.160 |

*Note.* Model 1 evaluated the effect of cost on discounting AUC values of delayed HIV vaccines while Model 2 assessed the impact of participant covariates. Statistically significant values are **bolded**.
\* $p < 0.05$ \*\* $p < 0.01$ \*\*\* $p < 0.001$

## 3 Experiment 2

The objective of Experiment 2 was to assess how vaccines are discounted under increasing levels of uncertainty when the vaccines are available for free or for $150, depending upon the condition. Similar to Experiment 1, participants were randomly assigned into two groups: one had the opportunity to receive an HIV vaccine at no cost, while the other had the option to purchase the vaccine for $150. Participants in each group completed a series of choices between a less effective HIV vaccine that was readily available and a more effective vaccine that was probabilistically available. The probability of obtaining the more effective vaccine varied across choices and ranged from 10% to 90%.

### 3.1 Participants

Upon completing Experiment 1, participants transitioned directly into Experiment 2.

### 3.2 Procedure

Participants were permitted to partake in a simulated HIV Vaccine Probability Discounting Task (HVPDT) where they were provided with the following instructions:

> Please read and consider the following. Imagine a situation in which two HIV vaccines are available for XX only at health centers, hospitals, and clinics in your city. HIV Vaccine C is 55% effective and is always available. HIV Vaccine D is 95% effective but is not always available due to high demand. You have the same income and savings as you do now, and vaccines are the only available options for protecting against HIV. You are aware of the risks associated with not getting vaccinated, and you understand the benefits of receiving the vaccine. The following questions ask about the likelihood of you booking an appointment to receive the vaccine, given various scenarios. Remember that there are no “right” or “wrong” answers. Please respond honestly, as if you were actually in this scenario.

The value of XX varied by the experimental condition that the participants were assigned to – whether HIV vaccines were available for free or for a $150 fee. Participants reported their preferences between a less effective but always available HIV vaccine C and an HIV vaccine D that was 95% effective but had varying probabilities of availability: 10%, 25%, 50%, 75%, and 90%.

Please note that demographic information and additional personal covariates were collected after the completion of this experiment, as described in the Measures section of Experiment 1.

### 3.3 Statistical Analyses

Prior to any data analyses, the five likelihood values were converted to odds-against values using the following equation:

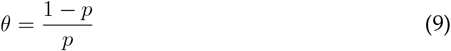

where *θ* is the odds against accessing the more effective HIV vaccine and *p* is the probability of not accessing the vaccine. The resulting values were: 0.11, 0.33, 1, 3, and 9. Consequently, the two-stage approach described in Experiment 1 was used to evaluate the discounting of both uncertain HIV vaccines and monetary rewards. Individual-specific discounting rates were estimated using the AUC method by normalizing both odds-against values (*θ*) and the value of the more effective HIV vaccine (*A*) so that they spanned from 0 to 1. AUC values were then estimated using the following:

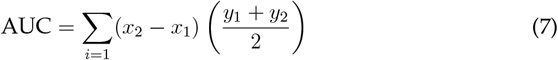

where *x*_1_ represents the normalized smaller odds against accessing the more effective HIV vaccine, *x*_2_ the normalized larger odds, and *y*_1_ and *y*_2_ are the corresponding normalized valuations of the more effective but uncertain HIV vaccine. At the aggregate level, the two-parameter hyperbolic discounting model was also used to describe probability discounting:

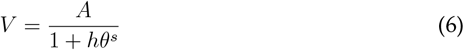

where *V* is the subjective value of the more effective HIV vaccine; *A* is the amount or magnitude of the vaccine; *θ* is the odds against accessing the more effective vaccine; a parameter, *h*, reflects the discounting rate, and another parameter, *s*, depicts sensitivity to increasing odds-against receiving the more effective vaccine.

As was the case in Experiment 1, the primary outcome variable was the AUC values for discounting uncertain HIV vaccines. Due to the non-normal distribution of AUC values, a logarithmic transformation was applied before proceeding to the second stage of analysis. This subsequent phase involved a multiple linear regression analysis conducted through three separate models. The first model used the experimental conditions as predictors to assess their direct impact on discounting AUC values. The second model examined the influence of demographic variables and other personal characteristics of participants. The third model combined both sets of predictors.

### 3.4 Results

Incomplete or missing entries were excluded, leading to the removal of 147 individuals, constituting 37.4% of the initial dataset. Consequently, data from 246 participants were carried forward for further analysis. Table 3 provides a summary of the demographic characteristics of study participants.

**Table 3.**
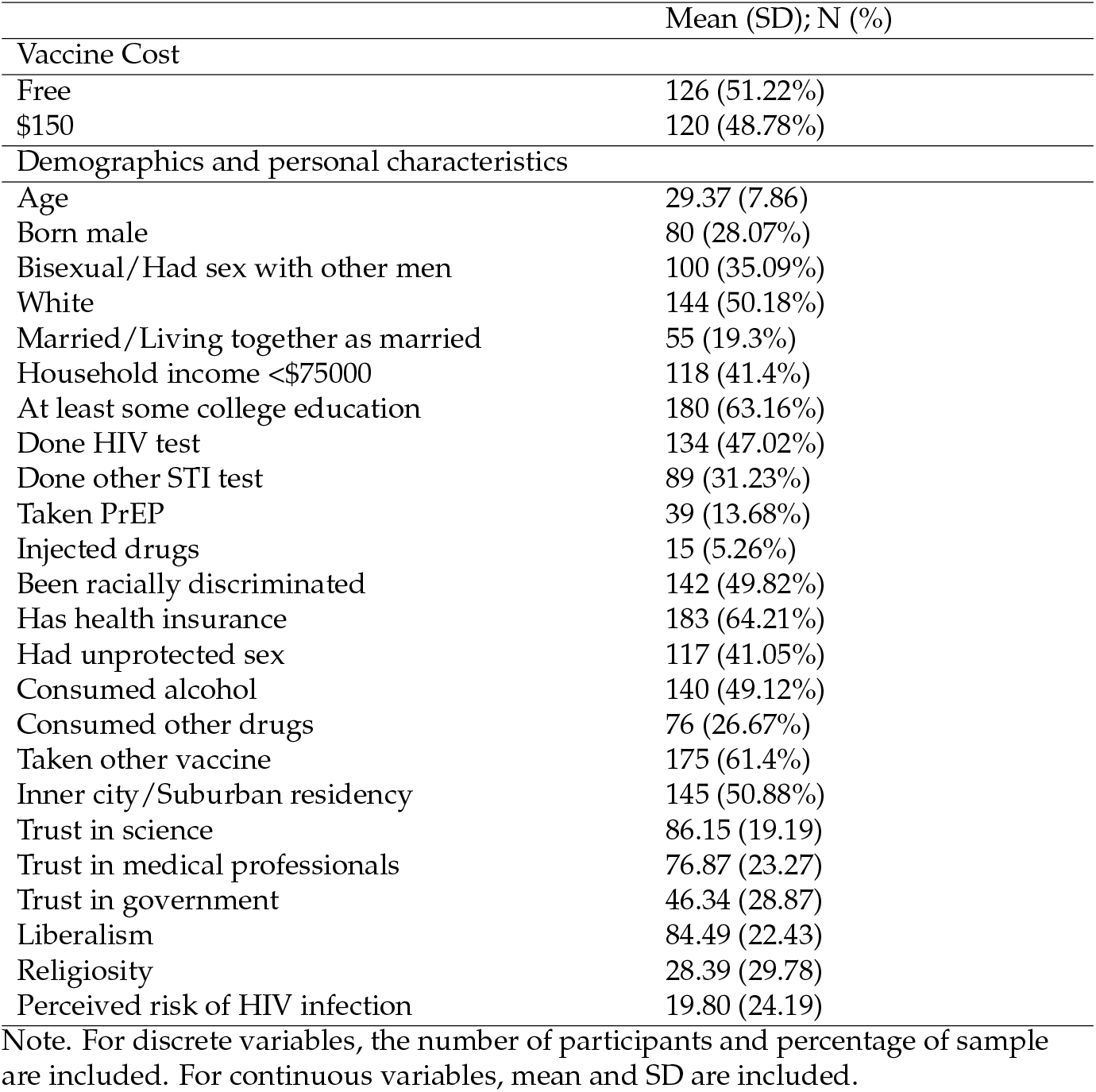
Characteristics of Participants Meeting Inclusion Criteria for Experiment 2 (N = 246)

|  | Mean (SD); N (%) |
| --- | --- |
| Vaccine Cost |  |
| Free | 126 (51.22%) |
| \$150 | 120 (48.78%) |
| Demographics and personal characteristics |  |
| Age | 29.37 (7.86) |
| Born male | 80 (28.07%) |
| Bisexual/Had sex with other men | 100 (35.09%) |
| White | 144 (50.18%) |
| Married/Living together as married | 55 (19.3%) |
| Household income <\$75000 | 118 (41.4%) |
| At least some college education | 180 (63.16%) |
| Done HIV test | 134 (47.02%) |
| Done other STI test | 89 (31.23%) |
| Taken PrEP | 39 (13.68%) |
| Injected drugs | 15 (5.26%) |
| Been racially discriminated | 142 (49.82%) |
| Has health insurance | 183 (64.21%) |
| Had unprotected sex | 117 (41.05%) |
| Consumed alcohol | 140 (49.12%) |
| Consumed other drugs | 76 (26.67%) |
| Taken other vaccine | 175 (61.4%) |
| Inner city/Suburban residency | 145 (50.88%) |
| Trust in science | 86.15 (19.19) |
| Trust in medical professionals | 76.87 (23.27) |
| Trust in government | 46.34 (28.87) |
| Liberalism | 84.49 (22.43) |
| Religiosity | 28.39 (29.78) |
| Perceived risk of HIV infection | 19.80 (24.19) |

Note. For discrete variables, the number of participants and percentage of sample are included. For continuous variables, mean and SD are included.

The hyperbolic discounting function was employed to model the probability discounting of HIV vaccines under both free and $150 fee conditions as shown by the R-squared values for the two experimental conditions (Free: *R*^2^ = 0.99; $150 fee: *R*^2^ = 0.95). The aggregate *h*-value calculated was 0.3663 for the free vaccine condition and 0.3731 for the vaccine with the $150 fee. Figure 3 illustrates the best-fitting hyperbolic discounting function for each condition. Additionally, there were no significant differences in the AUC between the two groups (Free: *M* = 0.64, *SD* = 0.40; $150 fee: *M* = 0.60, *SD* = 0.39; *t*(244) = − 0.75, *p* = 0.45), as shown in a bar chart in Figure 4. Furthermore, upon investigating the correlation between the log-transformed AUCs from the Monetary Probability Discounting Task and the HIV Vaccine Probability Discounting Task, it was determined to be insignificant, with a correlation coefficient of *r*(185) = − 0.07 and a *p*-value of 0.34. Figure 3 displays the aggregate discounting curves for uncertain HIV vaccines and probabilistic monetary rewards. At the aggregate level, Figure 3 illustrates how as the odds against obtaining a more effective vaccine increase, the demand for the less effective vaccine rises accordingly – indicating a substitutionary relationship between the two vaccines. This also shows that the odds against receiving the more effective vaccine can function similarly to a cost/price variable in impacting vaccine demand.

**Figure 3.**
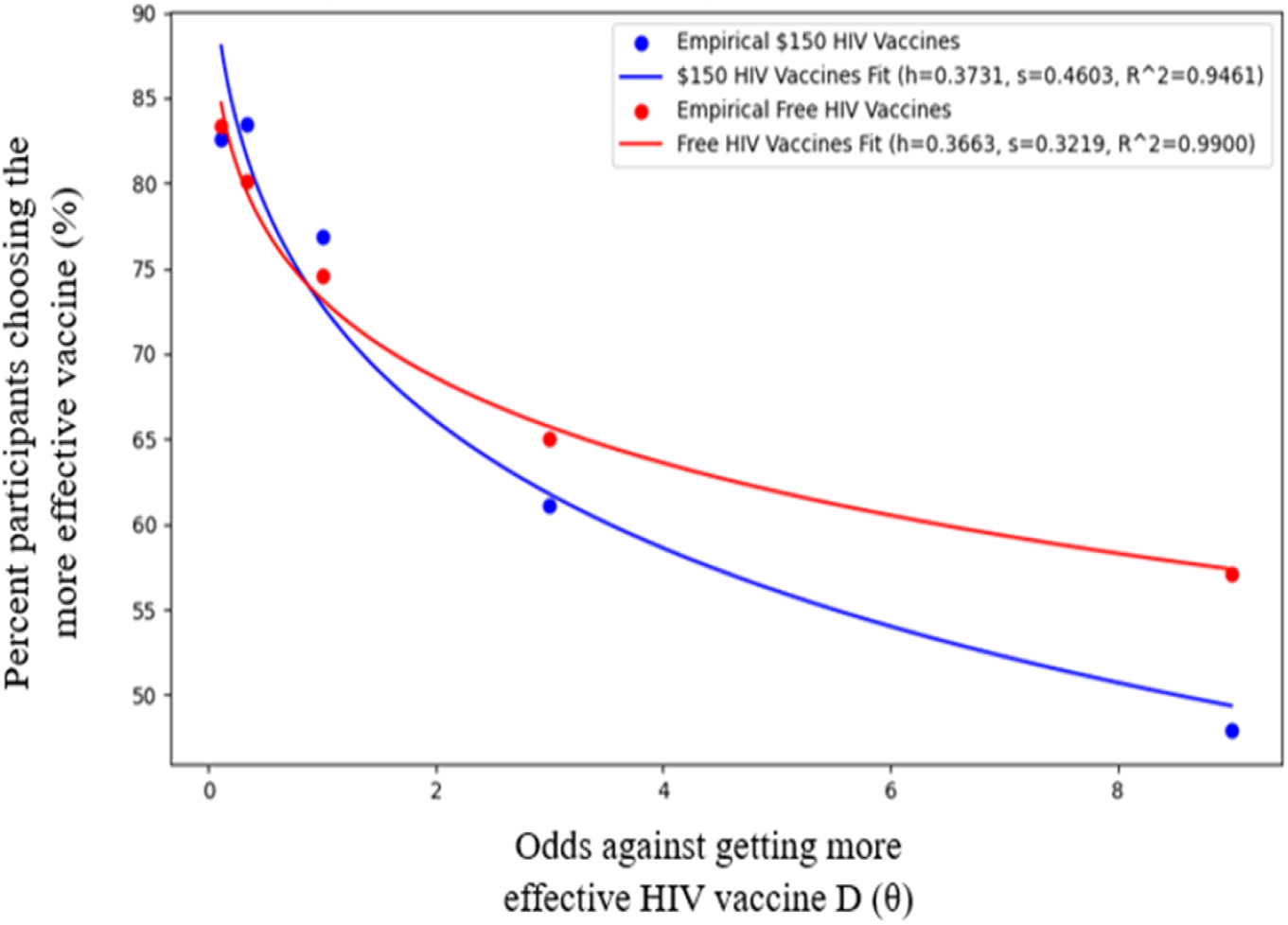
Aggregate Probability Discounting of a More Effective but Uncertain HIV Vaccine Available for a Fee vs Free. *Note*. Plotted are group discounting curves for a more effective but uncertain HIV vaccines available for free (red) and for a $150 fee (blue). Curves were plotted using a two-parameter hyperboloid model including a nonlinear scaling parameter on odds against getting the more effective vaccine. Note how at small odds against accessing the more effective vaccine, discounting of free vaccines is more pronounced than that of the $150 vaccines. At larger odds, however, discounting of the $150 vaccines is more than that of the free vaccines.

**Figure 4.**
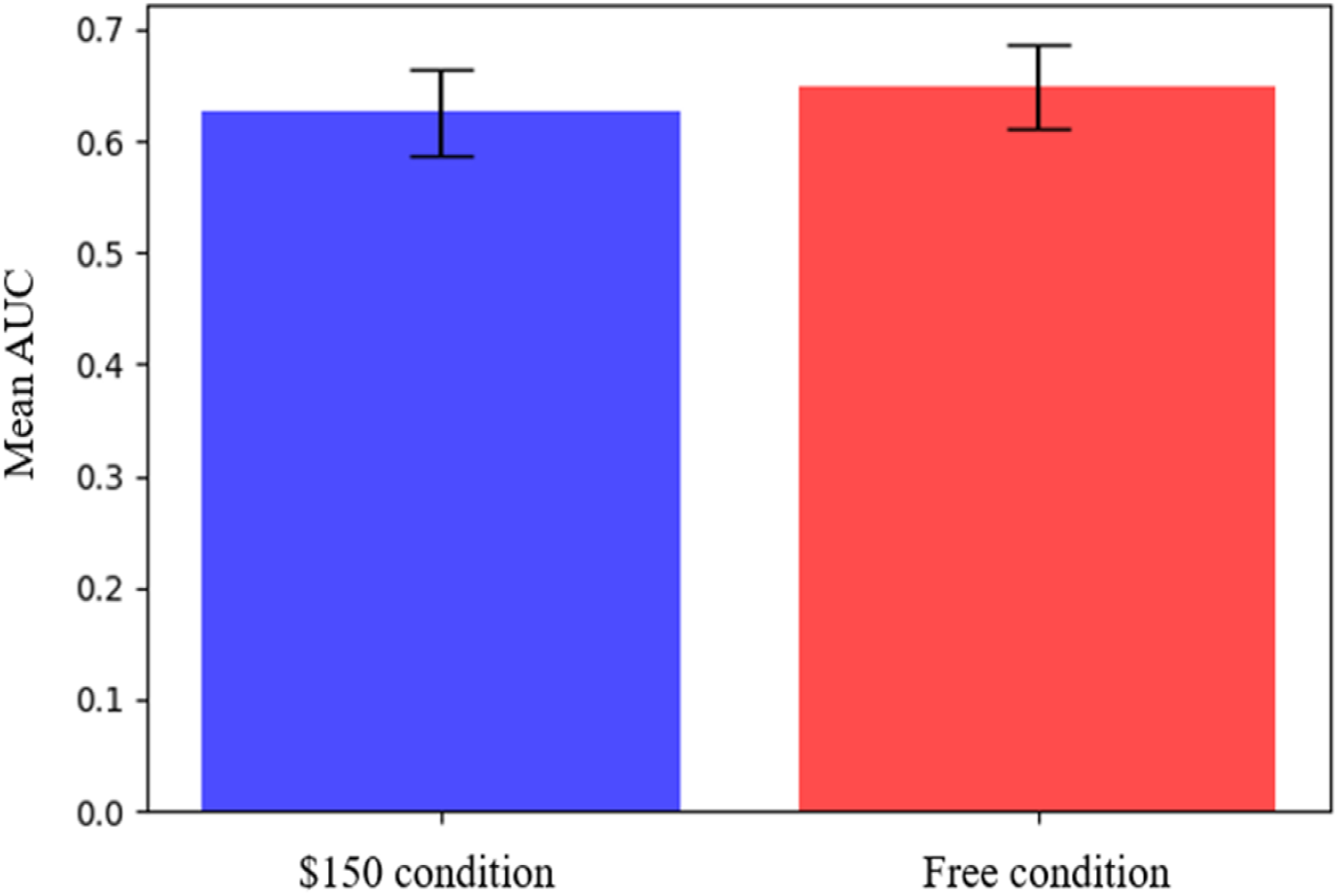
Bar Charts of Mean Probability Discounting AUCs for a More Effective but Uncertain HIV Vaccine Available for a Fee vs for Free. *Note*. The bars represent the group mean AUCs for a more effective but uncertain HIV vaccine available for free (red) and for a $150 fee (blue). Error bars indicate 95% confidence intervals.

Table 4 presents the outcomes of three regression models that explored what influences the variation in the log-transformed probability discounting AUCs. The initial model, which used the experimental condition of HIV vaccine costs (either free or at a cost of $150) as predictors, indicated a small effect of this context on probability discounting (*R*^2^ = 0.005). There were also no significant differences between the reference comparison group (vaccines for free) and the other condition (vaccine for $150) – even though the mean AUC was greater for the reference group. In the second model, the significant predictors for probability discounting included identifying as gay or bisexual (*β* = 0.18, *p <* 0.05), having health insurance (*β* = 0.33, *p <* 0.05), a history of undergoing screening tests for STIs and other than HIV (*β* = 0.18, *p <* 0.05), being politically left-leaning (*β* = − 0.004, *p* = 0.05), and age (*β* = 0.01, *p <* 0.05). The third model, which combined the experimental context with personal factors, showed a slight improvement in explanatory power compared to the second model (*R*^2^ = 0.198).

**Table 4.**
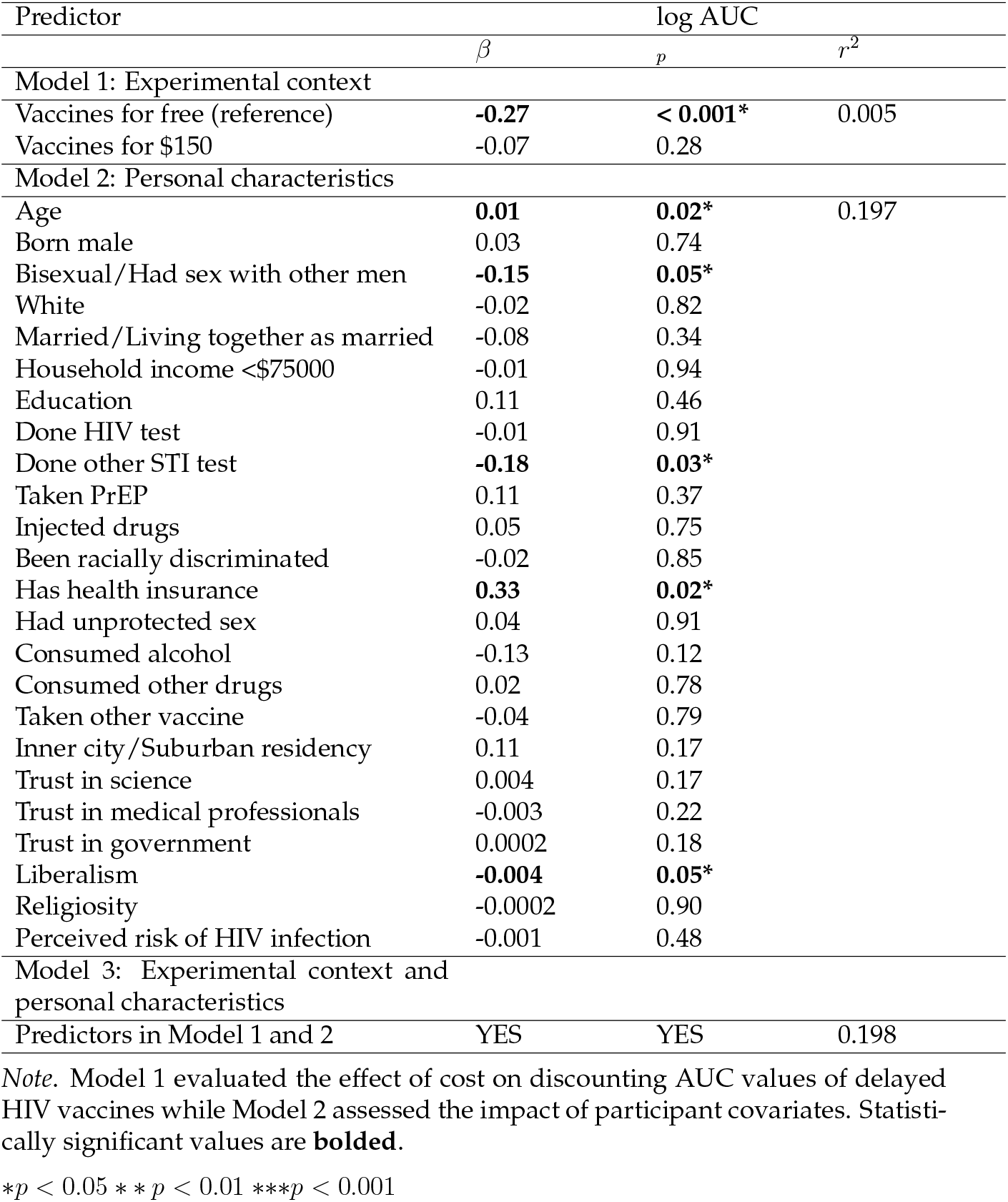
Regression Results Showing Effect of the Experimental Context of HIV Vaccines Available either for Free or at a $150 Fee and Personal Characteristics on Probability Discounting AUCs.

## 4 General Discussion

This research focused on investigating the influence of the monetary cost of the HIV vaccine on the extent of discounts due to delays or uncertainties in the availability of the vaccine. Experiment 1 sought to examine the impact of the cost of delayed vaccines – available either for free or for $150 – on how their value was discounted while experiment 2 assessed the rate of discount under increasing levels of uncertainty when the vaccines are available either for free or at a $150 cost based on several conditions. Both experiments had similar R2 values for both the delayed and probability discounting of both the free and paid vaccines, revealing similar discounting rates in both experiments.

The study indicated that the average AUCs for the HIV vaccine priced at $150 were less than those for the vaccine offered at no cost. This suggests that the free HIV vaccines underwent less significant discounting as uncertainty about accessing them rose, compared to those with a set price of $150. Participants in the free vaccine group exhibited less risk aversion than those in the $150 vaccine group. If participants in the $150 group viewed the cost as a minor loss, these results are inconsistent with Estle et al. (2006), who reported that probabilistic gains are discounted more steeply than probabilistic losses. That said, the effect size of the experimental context, where HIV vaccines were either free or had a $150 fee, was relatively small.

Participant age was associated with higher AUC values, indicating a lower likelihood of discounting the value of HIV vaccines under conditions of uncertainty. This result aligns with Olson et al. (2007) and Scheres et al. (2006), who found that delay discounting decreases with age. This could be because older adults tend to be less impulsive in their decision-making, allowing them to wait longer than younger adults, or because of experiences with delay as one age (Green et al., 1996; Olson et al., 2007). Additionally, socioeconomic factors may influence why delay discounting decreases with age, as age and income can affect the impulsivity of decision-making and willingness to accept delay (Green et al., 1996). However, this contrasts with Mok et al. (2020), who reported no relationship between age and delay discounting in monetary decisions, and Seaman et al. (2016), which observed higher rates of discounting for health benefits among older people.

A history of testing for sexually transmitted infections (STIs) was associated with higher AUC values, indicating a lower tendency to discount the value of uncertain HIV vaccines. This might be due to the perceived vulnerability to contracting these infections, which may lead to an increased interest in non-condom protective strategies, such as planning to avoid unprotected sex or using prophylactic drugs (Cannon & Celum, 2023; Thompson et al., 2021). This focus on protection may, in turn, explain why individuals with a history of STI testing may exhibit a more proactive attitude toward safeguarding their health (National Academies of Sciences, Engineering, and Medicine, 2021), including a greater willingness to receive novel HIV vaccines, even in the face of uncertainty. This finding is consistent with Martin-Smith et al. (2018), who found that heightened perceived susceptibility to STIs predicted STI testing, and Thompson et al. (2021), who observed that individuals engaging in risky sexual behaviors were more likely to have been tested for STIs in the past 12 months.

Participants with health insurance exhibited higher AUCs compared to those without insurance, suggesting insured individuals were more tolerant of uncertainties in accessing HIV vaccines. The higher likelihood of vaccination among insured individuals mirrors findings from Akpalu et al. (2020) and Getachew et al. (2023). This observation aligns with the notion that health insurance acts as a psychological and financial buffer (Finkelstein et al., 2018). Insured individuals may feel more secure in managing potential health challenges, including uncertainties associated with novel treatments. The higher AUCs observed in insured participants may also relate to the concept of moral hazard, where individuals tend to utilize more healthcare services, even when not entirely necessary, due to the reduced financial burden associated with health insurance coverage (Einav & Finkelstein, 2018). Identifying as gay or bisexual was linked with a lower probability AUC, indicating a lower tolerance for uncertainties in accessing HIV vaccines among this group. This could be due to their perceived higher risk of HIV infection. McGrath et al. (2019) found that among men who have sex with men (MSM), having more than four male partners in the last 12 months predicted accepting an HPV vaccine. Nadarzynski et al. (2018) reported perceived risk of infection predicted vaccine acceptance among MSM in the UK. This suggests gay and bisexual participants in the present study may prefer preventive measures they are certain to access.

Participants with a left-leaning political orientation exhibited a lower probability of AUC compared to other participants, suggesting they were less tolerant of uncertainties in accessing HIV vaccines. This is counterintuitive given that numerous studies, including Peng (2022), Lin et al. (2020), and Rabinowitz et al. (2016), indicated that left-leaning individuals are generally more positively disposed toward vaccine acceptance. However, this paradox can be understood in the context of left-leaning values prioritizing health equity and access to healthcare (Bodenheimer, 2005). Results from the study also showed that person-level factors such as race, HIV testing history, PrEP use, alcohol consumption, urban/inner city residency, and trust in medical professionals did not affect vaccination intentions. This could be attributed to the sampling method not adequately representing these key demographics.

Other personal factors did not significantly affect the probability discounting of HIV vaccines. Combining the financial scenarios of the HIV vaccines (free or $150 fee) with personal factors resulted in only a minor enhancement in explanatory power. This suggests that while both sets of variables are important, the pricing of the HIV vaccines might have a less substantial influence on how uncertain HIV vaccines are discounted. This observation contrasts with previous research findings, such as Mizak et al. (2021), who reported that for smaller amounts of money, losses were discounted more slowly than gains. The present studies’ findings also contrast with Weatherly and Derenne (2013), who found that delayed losses were discounted more slowly than delayed gains. These contrasts suggest that the $150 cost influenced participants’ discounting differently than in scenarios involving merely the gain of an HIV vaccine. This needs to be explored further in future studies using a wider range of costs.

The implication of these findings is important in policy formulation and public health interventions. Despite this, HIV vaccine uptake among vulnerable populations can be enhanced by leveraging the observed intolerance for delays and uncertainties to structure vaccine costs and delivery timelines strategically. This will help boost HIV immunity in the long run among vulnerable populations. This present research utilized Simulated Purchase Tasks (SPTs) as a tool to glean insights from both laboratory and clinical/field research to construct experimental simulations for potential policies and public health interventions. SPTs can simulate market conditions and potential vaccine intake without the financial and ethical rigors of direct clinical trials or premature market introductions. SPTs may offer valuable insights into people’s behaviors and preferences, such as the acceptance of novel health interventions like the HIV vaccine, because they are not conventional yes-or-no questions, which could lead to overestimation or underestimation.

Since the acceptance of novel health interventions like the HIV vaccine is becoming a crucial public health consideration, this study offers several significant contributions. One is by being one of the first studies to model HIV vaccine acceptance using behavioral economic methodologies to simulate decision-making in contexts where individuals lack prior experience. This research examines the nuanced decision-making processes associated with adopting a novel health intervention and requires participants to rely only on generalized decision-making strategies. The use of behavioral economics in recent years has demonstrated the relevance of simulations in predicting real-world behaviors, even in unfamiliar contexts. For instance, Wongsomboon and Webster (2023) employed delay discounting to model healthcare wait times, showing that participants’ valuation of delayed diagnostic tests varied by perceived disease severity (HIV versus chlamydia/gonorrhea). By extending such methodologies to novel health commodities like HIV vaccines, this study enhances the existing literature and underscores the utility of simulated purchase tasks in forecasting public health behavior.

Another strength of this research lies in its robust data collection from a population noted by Guillory et al. (2018) to be traditionally challenging to engage. Behavioral economic studies frequently recruit participants via crowdsourcing platforms like Amazon Mechanical Turk (MTurk); however, concerns about data quality on MTurk have risen in recent years. These issues include fraudulent responses from inattentive or inauthentic participants, often international users pretending to be Americans (Moss et al., 2021; Hauser et al., 2023), or responses generated by automated bots (Stokel-Walker, 2018; Kennedy et al., 2020; Storozuk et al., 2020), or familiarity with Simulated Purchase Tasks (SPTs) due to repeated exposure to experimental tasks, potentially biasing responses (Strickland & Stoops, 2019). To address these challenges, this study used alternative recruitment strategies, engaging participants through Reddit and community-based organizations targeting the SGMs community. While this method has its limitations, such as potential selection bias from self-motivated participants, it offers higher-quality data. Participants recruited through these channels are more likely to be actively engaged, resulting in more thoughtful and accurate responses.

Although the present study offers valuable insights, several limitations should be considered when interpreting its findings. A key limitation is the use of crowdsourcing for participant recruitment. As a non-probability convenience sampling method, crowdsourcing is prone to selection bias. It often relies on volunteers who frequently participate in multiple online panels, which can undermine the validity of the results and limit the generalizability of the study’s conclusions (Cornesse et al., 2020).

An additional limitation of the present study is the lack of information about the HIV status of participants, which could influence the perceived value and acceptance of preventative HIV vaccines. Without data on HIV status, the study could not examine differences in demand for and discounting of HIV vaccines attributable to this factor. Future studies will benefit from comparing the behavioral economic indices in HIV-positive and HIV-negative individuals.

The study’s design, where all participants engaged with the simulated tasks in a consistent order, presents a notable limitation (Gama et al., 2009). This uniform sequencing could inadvertently affect responses and requires further investigation to understand its impact fully. Additionally, the singular session format for data collection introduces the risk of participant fatigue, potentially leading to higher dropout rates and biasing the study’s outcomes (Olson, 2014). This fatigue could distort responses or reduce engagement with later tasks, affecting the reliability of the collected data.

Finally, there are several promising avenues for future research that could extend the understanding and applicability of these results. Firstly, future studies should strive to include a wider array of vulnerable population groups beyond the SGMs community, such as sex workers, people who inject drugs (PWIDs), and low-income communities. Including these groups would provide a more comprehensive view of vaccine acceptance and address potential disparities in access and willingness that may be specific to different communities. Next, future research could expand on methods to assess the substitutability and complementarity of HIV vaccines with other health interventions. This would offer valuable information on how bundled health commodities might influence vaccine acceptance and the potential synergistic effects that could enhance public health campaigns. Exploring additional conditions within delay and probability discounting tasks, such as manipulating price more parametrically using multiple price points, could reveal more nuanced understandings of how cost impacts vaccine valuation over time or under uncertainty. This would also allow for a more detailed analysis of price sensitivity and its role in vaccine acceptance.

## Data Availability

All data produced in the present study are available upon reasonable request to the authors

